# Rare and Common Germline and Somatic Variants Shape Immune Cytopenia Risk and Enable Risk Stratification

**DOI:** 10.64898/2026.08.26.26361484

**Authors:** Stennio Da Silva Faria, Julie Bineau, Rafael Moisan, Marc-André Legault, Estelle Lecluze, Thomas Pincez

## Abstract

The genetic risk factors of immune cytopenias are unclear. Immune cytopenias have been reported in various genetic contexts: 1) inherited error of immunity genes, mainly due to rare germline variants, 2) systemic lupus erythematosus, associated with common germline variants, 3) hematological malignancies, and 4) clonal hematopoiesis, the latter two due to somatic variants. However, the respective contribution and interaction of these variants remain to be investigated. Here, we used two large biobanks with whole genome sequencing data to systematically investigate the genetic contribution to immune cytopenia. We found that the four types of genetic variants independently contribute to immune cytopenia risk. We notably found that carriers of variants in some autosomal recessive genes of inherited error of immunity had an increased risk of immune cytopenia. Additionally, common variant-mediated risk of systemic lupus erythematosus also increased the risk of immune cytopenia. Overall, a third to a half of patients with immune cytopenia carried at least one of the four genetic risk variants investigated. Combining the four variants allowed stratifying the risk of immune cytopenia in both general and high-risk population. In general population, the 10- year incidence of immune cytopenia in the lowest and highest risk groups was 0.8‰ and 15‰, respectively. In sum, this work identified that different genetic risk factors can lead to immune cytopenia. A large proportion of individuals with immune cytopenia carried an underlying genetic risk factor. Finally, combining these genetic risk factors enabled risk stratification.

## Introduction

Immune cytopenias are the immune mediated destruction of platelets in immune thrombocytopenia (ITP) or red blood cell in autoimmune hemolytic anemia (AIHA).^1,2^ Disease evolution is highly variable, ranging from spontaneous remission to severe hemorrhage and life-threatening anemia.^3^ The genetic contribution to the risk of immune cytopenia is unclear. In a subset of cases, immune cytopenias can be secondary to a disease with known genetic component, mainly inborn errors of immunity (IEI), systemic lupus erythematosus (SLE), hematological malignancies, and clonal hematopoiesis.^4–6^ IEI (also called primary immunodeficiency), are monogenic diseases due to rare germline variants.^7^ Most of patients with immune cytopenias are not diagnosed with an IEI but the contribution of the rare variants in IEI genes to AIC is unknown. Moreover, ∼60% of IEI have an autosomal recessive (AR) inheritance but whether carrying a monoallelic variant in such gene increase the risk of immune cytopenias is unclear.^8^ SLE is a complex autoimmune disease that can affect virtually every organ.^9^ In adults, the risk of SLE is mediated by common germline variants which have been summarized in polygenic scores (PGS).^10^ Whether the common variant-mediated risk of SLE is also associated with immune cytopenias has not been investigated. Finally, hematological malignancies and clonal hematopoiesis are due to somatic variants in different genes and cell types.^11^ Both have been associated with an increased risk of immune cytopenia.^12–14^ Whether this risk is affected by germline variants is unknown.

The respective contribution of these four types of genetic variants to immune cytopenia has not been investigated. Their simultaneous investigation would provide unprecedented insights in immune cytopenias. Here, we leveraged the large UK Biobank (UKB) and All of Us (AoU) Research Program. We showed that joint analysis of these variants disentangles their respective contribution and identified that a substantial proportion of patients with immune cytopenia carry a genetic risk factor. Combination of genetic variants allow risk stratification of immune cytopenias in both general and high-risk populations.

## Methods

### Population

We analyzed genomics data from whole-exome sequencing dataset of the UKB (n=493,659 individuals) and the exome subset of whole genome sequencing dataset from the AoU Research program v8 (n=414,830 individuals).^15,16^ As suggested by previous research, we restricted our included AoU participants recruited on EHR sites to focus on participants with reliable EHR linkage (n=270,634 individuals).^17^ Mean (± standard deviation [SD]) age at genetic testing and last follow-up was 56.24 (± 8.09) and 72.70 (± 8.10) years for UKB and 53.32 (± 16.75) and 56.60 (± 16.72) years for AoU, respectively.

Patients with immune cytopenia were identified using ICD10 code for UKB (“D69.3 Idiopathic thrombocytopenic purpura” for ITP and “D59.1 Other autoimmune haemolytic anaemias” for AIHA) and Observational Medical Outcomes Partnership (OMOP) Common Data Model concept for AoU. OMOP is a common data model used by AoU to map source diagnosis codes to standardized clinical concept identifiers. It harmonizes different phenotype definitions that are used across the various health systems contributing to AoU, which use different systems.

We identified 937 patients with ITP (mean age 64.35 ± 10.55 years) and 251 patients with AIHA (mean age 66.39 ± 9.53) in the UKB (24 patients had both ITP and AIHA). In AoU, we identified 802 patients with ITP (mean age 56.13 ± 17.03 years) and 236 with AIHA (mean age 57.72 ± 17.41 years), in which 22 patients had both ITP and AIHA.

We also identified patients with SLE and hematological malignancies (**Supplemental Table 1**).

### Clonal hematopoiesis

In the UKB, we used the available expert-curated list of clonal hematopoiesis published by Vlasschaert et al.^18^ Clonal hematopoiesis was analyzed from the whole exome sequencing of 454,787 participants using the blood sample obtained at inclusion. The pipeline included 58 known clonal hematopoiesis driver genes (after excluding genes not associated with age) and was restricted to variants with variant allele frequency (VAF) ≥2%, corresponding to the threshold of clonal hematopoiesis of indeterminate potential.^19^ Clonal hematopoiesis calling was based on Mutect2 and removed variants with low coverage, sequencing artefacts, and germline variants.^20^

Importantly, some large clone with VAF near 50% can be considered as germline variants and no other tissue than blood is available in the UKB to discriminate the two types of variants. This is especially the case for *TET2*, in which variants in the catalytic domain can exist both at germline and somatic states. To limit this classification error, the authors performed a binomial test with age at analysis to detect specific variants more likely reflect clonal hematopoiesis rather than germline variants.

### Rare variants analysis

We filtered the germline variants in a subset of 62 IEI genes: those previously associated with ITP and/or AIHA using a recently curated list.^21^

In UKB, we used the variant annotation results provided by UKB to select the LoF variants,^15^ and annotated likely deleterious missense variants using REVEL and AlphaMissense based on the Ensembl Variant Effect Predictor (VEP) plugin.^22,23^ Then, we used plink v2.00a5.8,^24^ to select variants with a minor allele frequency (MAF) <0.1% or <1% using frequency in the whole cohort analyzed.

In AoU, we used a different annotation tool to validate the robustness of our findings. We selected LoF variants based on GnomAD annotations (“splice_acceptor_variant”, “splice_donor_variant”, “stop_gained”, “frameshift_variant”, “start_lost”) and filter on the allele frequency.^25,26^ We considered the variant burden by clumping all LoF and/or likely deleterious missense variants in the 62 gene set.

As mentioned earlier, variants in some genes, especially *TET2*, can correspond to somatic clonal hematopoiesis variants rather than germline variants. To remove the variants that may reflect clonal hematopoiesis,^20^ we took into account the results from Vlasschaert et al.^18^ Briefly, we filtered out variants that we selected in our rare LoF/missense variants filtering steps but were also identified as CHIP somatic variants by Vlasschaert et al.^18^

### PGS for SLE

We used three published weighted PGS for SLE: 1) the standard PGS generated in the UKB and which weights are not available for external replication,^27^ 2) the PGS000196,^28^ and 3) the PGS0004917 in AoU.^29^ PGS000196 and PGS0004917 scoring sheets with allele weights were obtained through the PGS Catalog,^30^ and the PGS computed in AoU using AoUPRS package.^31^ We used OptSurvCutR (genetic method based on rgenoud package and maximum log rank) to identify the cutpoints of PGS_SLE_ in the UK Biobank.^32,33^ The cutpoint identified was 3.03 z-scores. We used the package pROC to compute receiver operating receiver curve (ROC).^34^

For the combination of PGS_SLE_ with rare and somatic variants, we defined high PGS_SLE_ as values >1.5 SD to obtain comparable risk between LoF and high PGS_SLE_ but performed sensitivity analyses using >1 and >2 SD.

### End of follow-up and censoring dates

Follow-up windows were defined separately in each cohort to reflect the different structure of their linked health records.

In UKB, censoring dates were constructed from four sources — the main participant table (which also provided cancer diagnosis dates), the hospital inpatient register (HESIN), the primary care (GP) records, and the death register (DEATH) — covering 501,936 participants. Administrative censoring dates differ by nation (England, Scotland, Wales) and register. For each register, the censoring date was assigned from the register’s own nation whenever the participant had at least one record in that register; otherwise, it was derived from a participant-level consensus nation. When the register-derived nations were discordant — most plausibly reflecting relocation — priority was given to the nation recorded in HESIN or GP, and, when neither was available, to the nation of recruitment. For the GP register, when the data provider was unknown, the earliest (most conservative) provider-specific date within the consensus nation was retained.

In AoU, end of follow-up was defined at the participant level from the linked OMOP tables (Controlled Data Repository v8, release C2024Q3R4), under a single program-wide administrative cutoff of 1 October 2023. A conservative definition was used, anchored on the end of the EHR/PPI observation period rather than on the most recent clinical activity of any kind, because the outcomes analysed here were ascertained from EHR diagnoses. For each participant, the end of follow-up was taken as the death date when a primary death record was available and as the end of the EHR/PPI observation period otherwise (obs_period_end_date, used as the best available proxy for the last data transfer by the recruiting site); this date was then capped at the administrative cutoff.

### Statistical analyses

We used logistic and Cox regression (using survival R package)^35^ to compute odds and hazard ratio, respectively. All multivariate analyses were adjusted on sex, age at last follow-up and the 10 first principal components. We used age at last follow-up to consider the variable duration of follow-up between individuals but previously performed sensitivity analyses considering age at inclusion and confirmed this did not impact our results. For analyses of incident cases, we used different strategies to define start point in the two cohorts because of the different structure of UKB and AoU. In UKB, we used the inclusion in the cohort (corresponding to the timepoint of clonal hematopoiesis analysis) as start point and removed all individuals with ITP and/or AIHA before inclusion in UKB. In AoU, we used the first EHR as startpoint.

We used R (version: 2025.5.1.513) and GraphPad Prism (version: 2025.5.1.513) to perform statistical analyses and create figures.^36^ All tests were two-sided. We used the Benjamini-Hochberg method to compute the false discovery rate (FDR) to consider multiple testing.^37^

## Results

### Rare variants in IEI-genes increase the risk of immune cytopenia

Although immune cytopenias can occur in the rare context of IEI, the contribution of IEI variants to immune cytopenia has not been systematically investigated. Moreover, heterozygous carriers have an increased risk of immune cytopenia remain unclear.

To assess the contribution of rare variants to immune cytopenia risk, we first analyzed rare (MAF <0.1%) predicted deleterious (LoF or REVEL ≥0.9) variants in 62 genes of IEI previously associated with ITP and/or AIHA.^21^ In a multivariate model adjusted for age, sex, and the first 10 principal components, we found that patients carrying such variants were more likely to develop ITP and AIHA (**Figure 1A** and **Supplemental Table 2**). This effect was driven by LoF variants, which were strongly associated with both immune cytopenia risk whereas deleterious missense variants were not. We confirmed our findings using a more liberal MAF threshold (<1%), with more liberal REVEL scores (≥0.5), and with another algorithm for identifying deleterious missense variants (AlphaMissense). We validated our findings for LoF in the AoU cohort using a different annotation tool (GnomAD). For all further analyses, we considered only LoF with MAF <0.1%.

**Figure 1.**
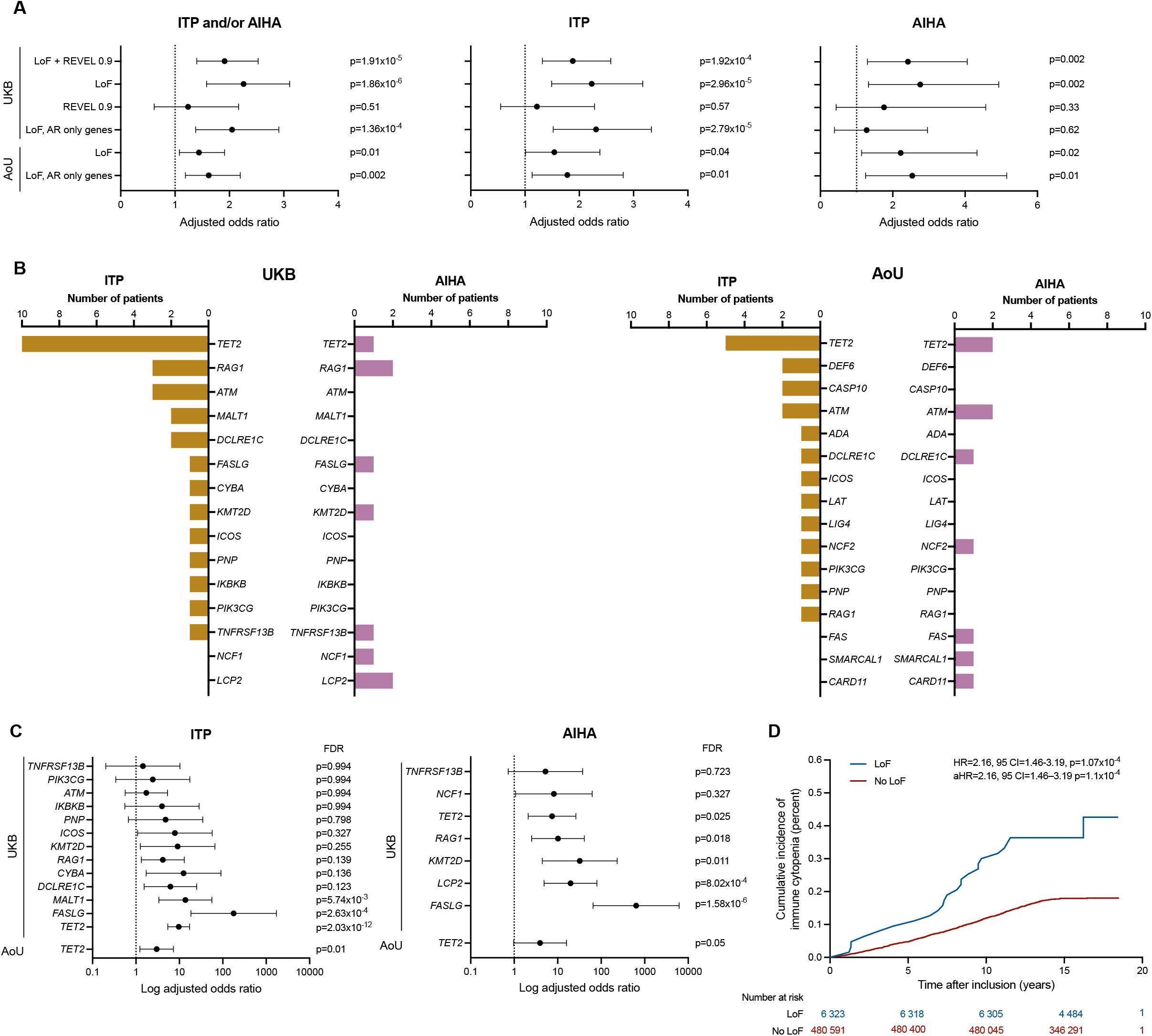
LoF variants in IEI genes increases the risk of immune cytopenia. A) Association between rare (MAF <0.1%) predicted deleterious variants and the risk of ITP and AIHA in the UKB and AoU using different criteria for deleteriousness. We considered variants in the 62 IEI genes (or the subset of 41 AR genes) previously associated with ITP and/or AIHA. B) Number of genes carrying a LoF variant in patients with ITP and AIHA in the UKB. C) By-gene analysis for the association of LoF variants and ITP and AIHA. Each gene was tested in a separate multivariate model. D) Cumulative incidence of immune cytopenia based on the presence of LoF variants in the UKB.

Among patients with immune cytopenia, we found variants in 15 genes in the UKB and 16 in AoU (**Figure 1B**). The most frequent rare variant-harboring gene was *TET2* in both cohort (n=11 in UKB and n=7 in AoU). In almost all cases (32/33 [96%] in the UKB and 28/29 [97%] in AoU), LoF were monoallelic. In both cohorts, 80% of the genes (which included 86% and 83% of the LoF variants in UKB and AoU cohorts, respectively) were classified as AR (biallelic) in the IUIS classification (considering *TNFRSF13B* as an autosomal dominant gene). By restricting our analysis to genes with AR inheritance, we found that carriers of heterozygous LoF in these genes had a higher risk of immune cytopenia in both UKB and AoU cohorts.

Burden analysis enables the joint analysis of related genes, potentially increasing power. Nevertheless, we sought to identify if LoF in specific genes were associated with immune cytopenia risk. We found that LoF in *TET2* and *FASLG* were individually associated with both ITP and AIHA in UKB (**Figure 1C**). We could replicate the association between finding in *TET2* and ITP in AoU. In UKB, *KMT2D* and *LCP2* were associated with AIHA and *MALT1* was associated with ITP but no patients with immune cytopenia carried variants in these genes in AoU (**Supplemental Table 3**).

Finally, we investigated the effect of LoF variant on the cumulative incidence of immune cytopenia after recruitment in the UKB using Cox regression model. We identified that patients carrying LoF variant had a 2.3-fold increased risk of immune cytopenia compared to those without (**Figure 1D**). Ten years after the inclusion in the UKB, the cumulative incidence of immune cytopenia in individuals with and without LoF was 0.33% and 0.12%, respectively.

In sum, our results suggest that LoF in IEI genes increases the risk of immune cytopenia. Specifically, carrying one LoF variant in an AR IEI gene is a risk factor for immune cytopenia.

### Genetically determined risk of SLE increases immune cytopenia risk

Some patients with immune cytopenia will develop SLE, and *vice versa*.^38,39^ SLE risk is influenced by common variants but whether these variants also influence immune cytopenia risk in unknown.

To investigate whether an increased genetic risk of SLE was also associated with immune cytopenia, we used published PGS for SLE (PGS_SLE_). We found that a higher PGS_SLE_ value was associated with both AIHA and ITP risk in the UKB (**Figure 2A** and **Supplemental Table 4**). We confirmed our findings in AoU using two different PGS_SLE_. This association was robust to adjusting for LoF variant burden. To ensure our findings were not confounded by patients with immune cytopenia having also developed a SLE, we perform a sensitivity analysis after having excluded these patients. We confirmed the association between PGS_SLE_ and ITP and AIHA in both the UKB and AoU cohorts.

**Figure 2.**
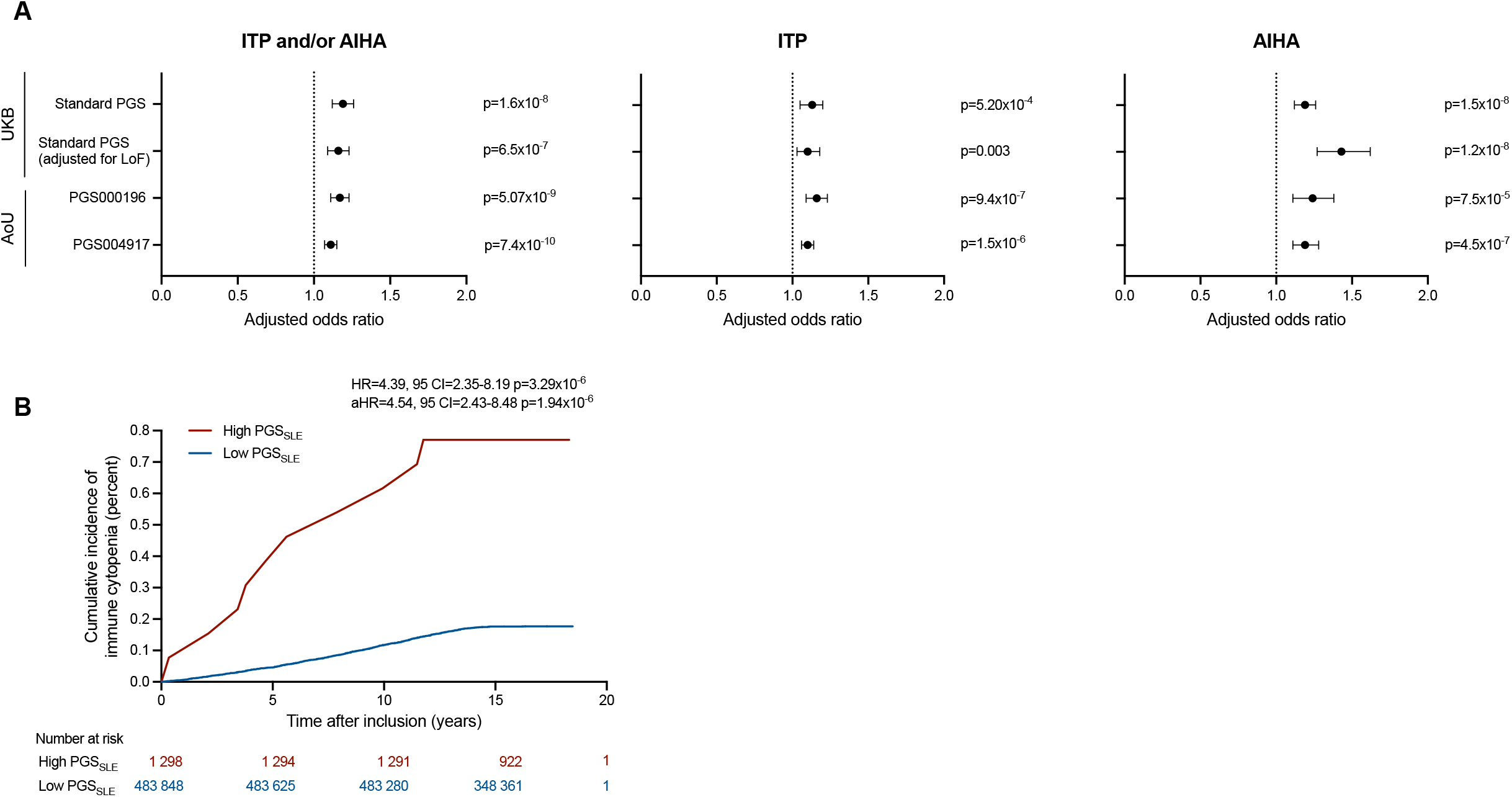
PGS_SLE_ allow stratifying the risk of immune cytopenia. A) Association between three PGS_SLE_ and the risk of ITP and AIHA in the UKB and AoU. The odds ratios are shown per SD of PGS_SLE_. B) Cumulative incidence of immune cytopenia in the general population based on the PGS_SLE_ in the UKB. The cut-off for high and low PGS_SLE_ groups was 3.03 z-scores.

We then assessed the cumulative incidence of immune cytopenia based on PGS_SLE_. In the UKB, we found a 4.4-fold increased risk of immune cytopenia in patients with high PGS_SLE_ value (**Figure 2B**). Ten years after the inclusion in the UKB, the cumulative incidence of immune cytopenia in individuals with a high and low PGS_SLE_ value was 0.62% and 0.12%, respectively.

In sum, our results suggest that a higher genetic risk of SLE also increases the risk of immune cytopenia.

### Rare variants in IEI genes, PGS_SLE_, and hematological malignancies independently contribute to the risk of immune cytopenia and allow risk stratification

Hematological malignancies are due to somatic variants and have been associated with immune cytopenia. However, the respective contribution and interaction between somatic variants and germline rare and common variants remain unexplored.

Using a multivariate model, we found that LoF in IEI genes, PGS_SLE_, and hematological malignancies were independently associated with ITP and AIHA risk (**Figure 3A** and **Supplemental Table 5**). In both UKB and AoU, the highest risk was carried by hematological malignancies and LoF and PGS_SLE_ were associated with a similar risk. We then reasoned that combining these variants would allow determining different risk groups. Thus, we compared the cumulative incidence of immune cytopenia among three genetically defined groups of individuals: those with hematological malignancies, those with LoF in IEI genes and/or high PGS_SLE_, and the others. In both UKB and AoU cohorts, we found these three genetic variant-based categories had different risk of immune cytopenia (**Figure 3B**). Ten years after the inclusion in the UKB, the cumulative incidence of immune cytopenia in individuals with hematological malignancy, those with LoF in IEI genes and/or high PGS_SLE_, and those without genetic variants were 15‰, 1.7‰, and 0.9‰, respectively. In the AoU, these numbers were 16‰, 3.9‰, and 2.8‰, respectively.

**Figure 3.**
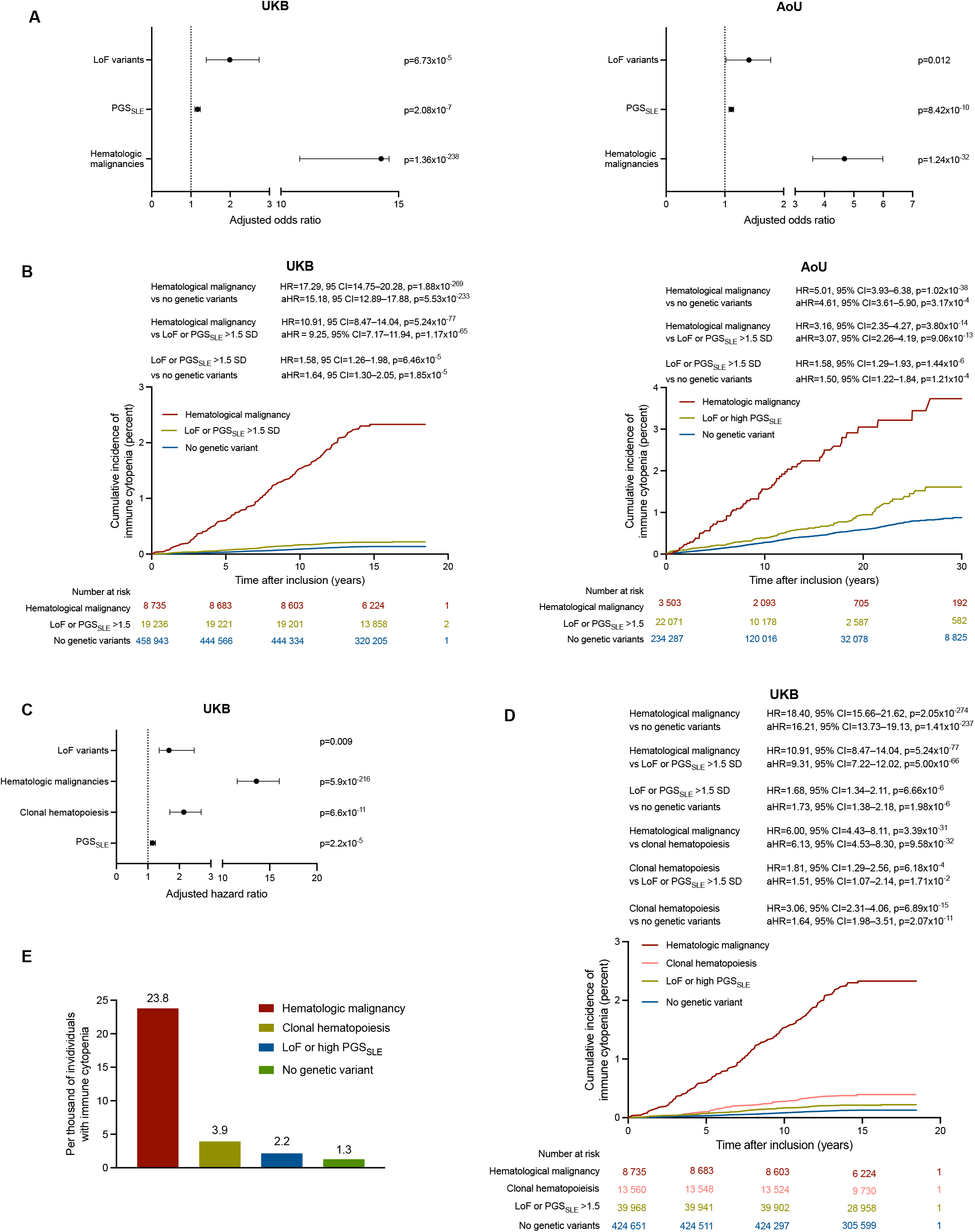
Stratification of immune cytopenia risk based on LoF variants, PGS_SLE_, hematological malignancy, and clonal hematopoiesis. A) Association between LoF variants, PGS_SLE_, and hematological malignancy with the risk of immune cytopenia in a multivariate logistic regression model. The odds ratios are shown per SD of PGS_SLE_. B) Cumulative incidence of immune cytopenia in UKB (left) and AoU (right) based on the three genetic variant-based groups: individuals with hematological malignancies, those with LoF in IEI genes and/or high PGS_SLE_, and the others. We considered PGS_SLE_ >1.5 SD as high PGS_SLE_ to obtain comparable risk between LoF and high PGS_SLE_ but found similar results using >1 and >2 SD (**Supplemental Table 7**). C) Association between LoF variants, PGS_SLE_, hematological malignancy, and clonal hematopoiesis with the risk of immune cytopenia in a multivariate logistic regression model. The odds ratios are shown per SD of PGS_SLE_. D) Cumulative incidence of immune cytopenia in UKB based on the four genetic variant-based groups: individuals with hematological malignancies, those with clonal hematopoeisis, those with LoF in IEI genes and/or high PGS_SLE_, and the others. We considered PGS_SLE_ >1.5 SD as high PGS_SLE_ but found similar results with >1 and >2 SD (**Supplemental Table 9**). E) Proportion of patients with immune cytopenia according to the genetic variant identified. All comparisons are statistically significant (p <4.7×10^−4^).

Clonal hematopoiesis, another category of somatic variant, has previously been associated with both ITP and AIHA. We investigated whether considering it allowed to refine our risk model. We investigated only the UKB only as the sample size of individuals with clonal hematopoiesis analysis was too low in AoU (n=98,560).

We found that clonal hematopoiesis, hematological malignancies, LoF in IEI genes and PGS_SLE_ were independently associated with immune cytopenia risk (**Figure 3C** and **Supplemental Table 6**). Clonal hematopoiesis was associated with an intermediate risk, between hematological malignancies and LoF in IEI genes and high PGS_SLE_ (**Supplemental Table 7**). We added clonal hematopoiesis as an additional genetically defined risk category to our previous categories for risk stratification. Ten years after the inclusion in the UKB, the cumulative incidence of immune cytopenia in individuals with LM, those with clonal hematopoiesis, those with LoF in IEI genes and/or high PGS_SLE_, and those without genetic variants were 15‰, 2.7‰, 1.7‰, and 0.8‰, respectively. (**Figure 3D** and **Supplemental Table 8**). We confirmed the difference between the four groups while considering the prevalence of immune cytopenia throughout all the follow-up available in the UKB (**Figure 3E**).

Finally, to assess whether some of these four genetic determinants modulate the risk of one other, we tested all two-term interactions in the UKB. We found a nominal interaction between clonal hematopoiesis and hematological malignancies (aHR=0.53, 95% CI=0.32- 0.87, p=0.01) while the other interaction terms were not significant (**Supplemental Table 9**).

In sum, the four genetic variants independently contribute to immune cytopenia risk. Their combination allows effective risk stratification with markedly risk difference between the groups, defining both high- and low-risk groups. While rare and common germline and somatic variants appear to carry an additive independent risk, our results suggest that the two types of somatic variants may not represent an additive risk.

### Genetic variants contribute to a substantial proportion of immune cytopenias

We analyzed the proportion of individuals with immune cytopenia who had an underlying genetic variant increasing the risk of immune cytopenia (**Figure 4A**). LoF variants were found in 2.8% of patients with immune cytopenia (vs 1.2% in the whole UKB). PGS_SLE_ >2 SD were found in 5.2% of patients with immune cytopenia (21.3% had PGS_SLE_ value >1 SD). Hematological malignancies having occurred before immune cytopenia was found in 27.2% of cases. Clonal hematopoiesis was detected before immune cytopenia in 8.5% of cases. Overall, 36.7% of individuals carried a genetic risk variant considering PGS_SLE_ >2 SD and 48.3% considering PGS_SLE_ >1 SD. In most cases (30.3%), only one genetic variant was found and only 6.4% of individuals with immune cytopenia carried ≥2 genetic variants that may have increased the risk (**Figure 4B**).

**Figure 4.**
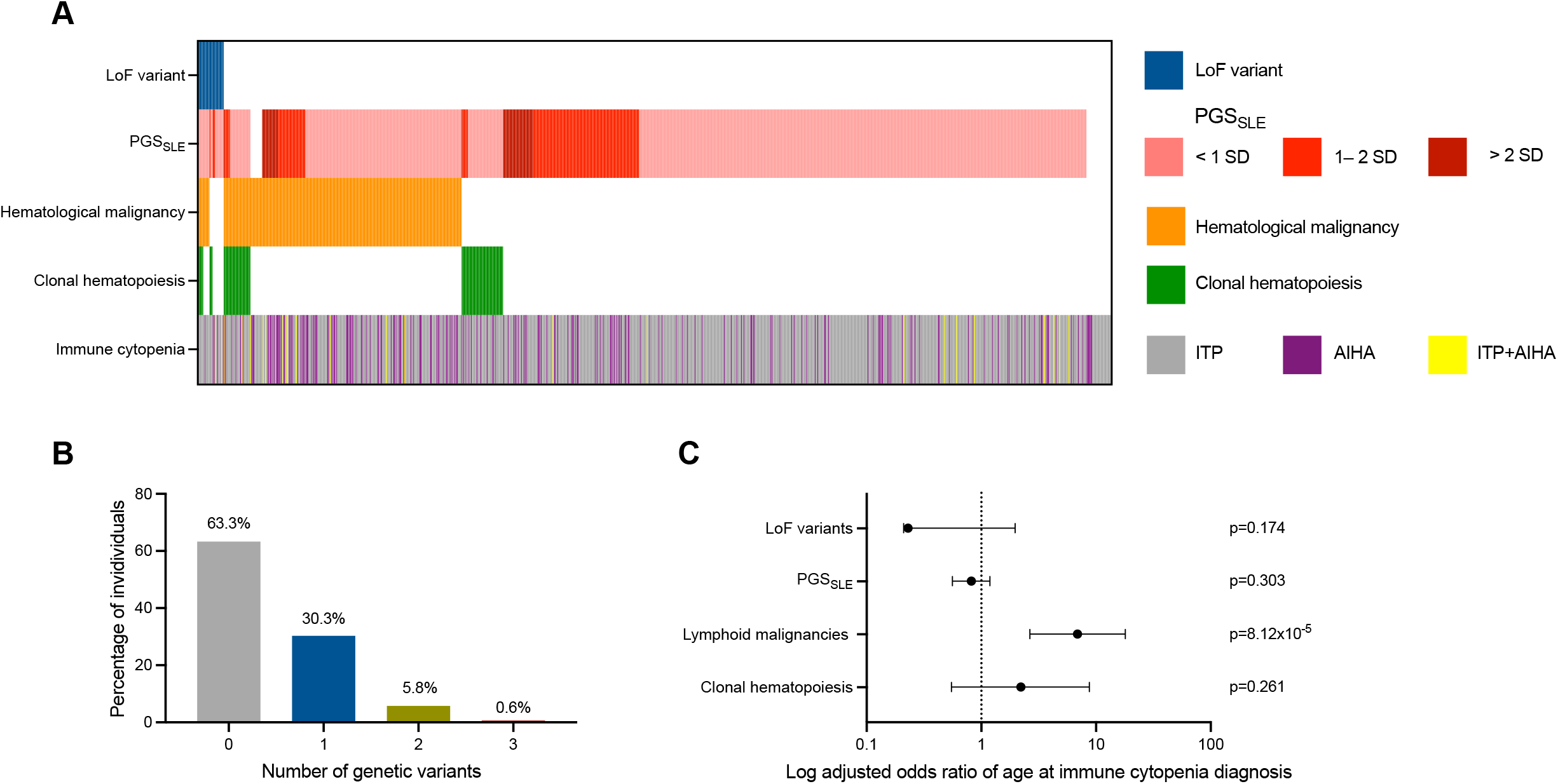
Proportion and co-occurrence of genetic variants contributing to immune cytopenia. A) Heatmap of genetic variants in the 1,164 individuals with immune cytopenia of the UKB. B) Number of genetic variants in individuals with immune cytopenia considering PGS_SLE_ >2 SD. C) Association of the four genetic variants with age at immune cytopenia diagnosis in a multivariate linear regression model.

We then analyzed the association of genetic variants with age at immune cytopenia diagnosis. Although germline variants tended to be associated with younger age at diagnosis and somatic variants with older age at diagnosis, only hematological malignancies had a significant association (**Figure 4C** and **Supplemental Table 10**).

Thus, one third to almost half of patients with immune cytopenia carried a genetic variant that increased their risk of immune cytopenia. The most frequent were somatic variants and most individuals carried only one variant.

### Stratifying the risk of immune cytopenia in high-risk populations using rare variants in IEI genes and PGS_SLE_

We have shown that individuals with hematological malignancies and clonal hematopoiesis present an increased risk of immune cytopenia independently of rare LoF variants in IEI genes and PGS_SLE_. We reasoned that using these two types of germline variants could allow stratifying the risk of immune cytopenia in the high-risk population with somatic variants, eventually allowing individualized management.

We first analyzed whether LoF and PGS_SLE_ were associated with immune cytopenia risk in patients with hematological malignancies or with clonal hematopoiesis (**Figure 5A** and **Supplemental Table 11**). We found an association with LoF and PGS_SLE_ in individuals with hematological malignancies and an association with LoF but not PGS_SLE_ in individuals with clonal hematopoiesis.

**Figure 5.**
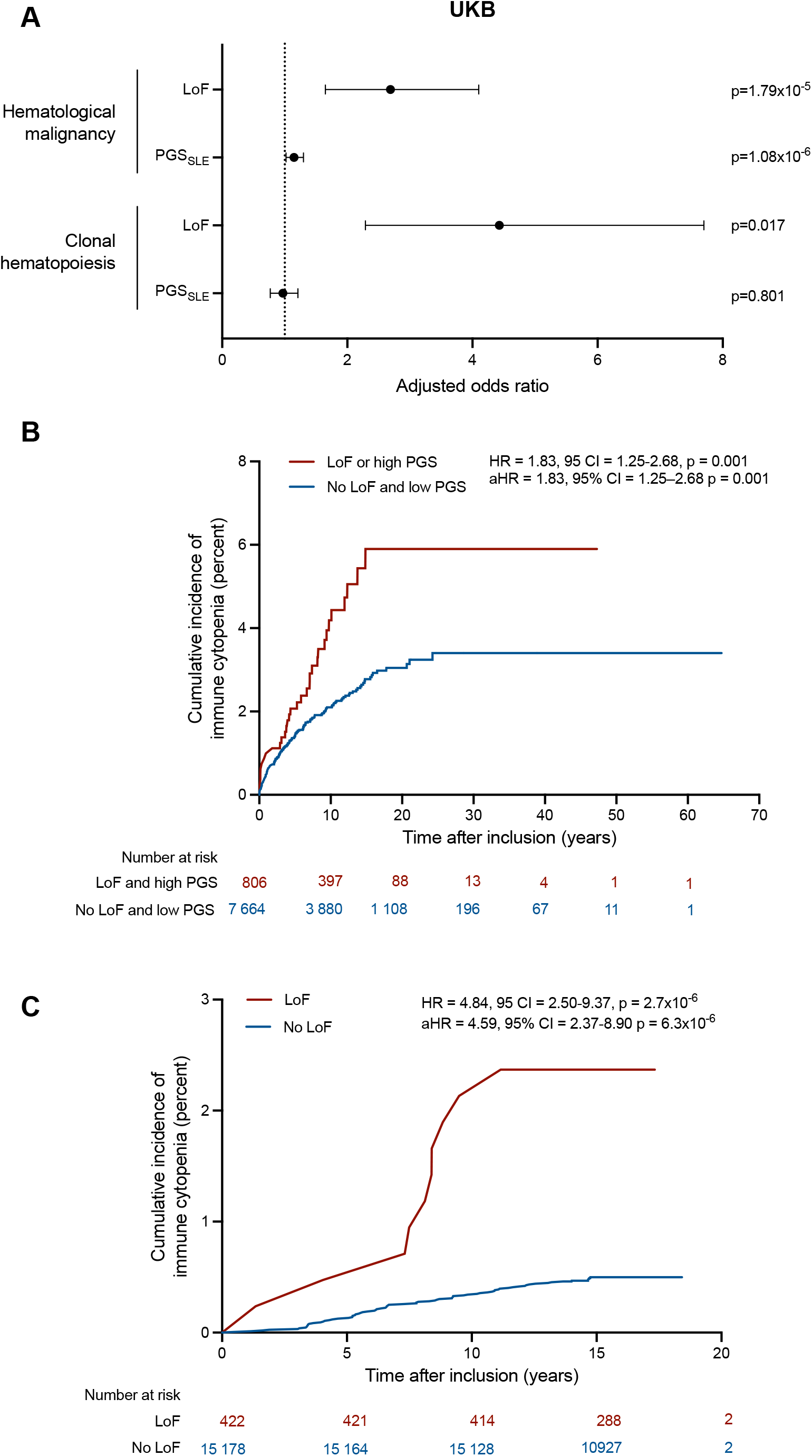
Stratification of immune cytopenia risk based on LoF variants in IEI genes and PGS_SLE_ in higher-risk populations. A) Association between LoF variants and PGS_SLE_ with the risk of immune cytopenia in individuals with hematological malignancy or clonal hematopoiesis. The odds ratios are shown per SD of PGS_SLE_. B) Cumulative incidence of immune cytopenia among patients with hematological malignancy based on the combination of LoF variants and PGS_SLE_ in the UKB. We considered has having a genetic variants individuals carrying a LoF or a PGS_SLE_ >1.5 SD. C) Cumulative incidence of immune cytopenia among patients with clonal hematopoiesis based on the presence of LoF variants in the UKB.

We then classified individuals with hematological malignancies according to the presence of LoF variant and PGS_SLE_ in two categories: those with either high PGS_SLE_ value or LoF variant and those with a low PGS_SLE_ value and without LoF (no individuals had both high PGS_SLE_ value and LoF). We found that individuals with LoF variant or high PGS_SLE_ value had a higher incidence of immune cytopenia (**Figure 5B** and **Supplemental Table 12**). Finally, we classified individuals with clonal hematopoiesis according to the presence of LoF. We found that individuals with LoF variant had a higher incidence of immune cytopenia (**Figure 5C**).

Thus, LoF and PGS_SLE_ allows stratification of immune cytopenia risk among two-high-risk population: individuals with hematological malignancies or clonal hematopoiesis.

## Discussion

Here, we showed that rare germline variants, common germinal variants, and two types of somatic variants, clonal hematopoiesis and hematological malignancies, are independent risk factors of immune cytopenia. Common germline variants also influenced the risk of SLE in individuals with immune cytopenia. We estimated that around one third to one half of individuals with immune cytopenia carried a contributing genetic variant. Finally, we showed that germline and somatic variants can be used for risk stratification of immune cytopenia in both general and higher-risk population.

Some IEI are associated with a very high risk of immune cytopenia.^40^ However, the large majority of patients with immune cytopenia are not diagnosed with an IEI in clinical setting. The proportion of individuals with immune cytopenia carrying an underlying IEI is unclear.^41^ Previous studies found variable results, likely depending on the population investigated and methodological approach.^4,42–46^ These studies investigated clinical-grade variants allowing to retain the diagnosis of IEI, which are mainly monogenic disorders with mendelian inheritance.^8^ One major and uninvestigated question is whether some variants in IEI genes may act as risk factors of immune cytopenia despite not leading to monogenic IEI. This question is especially relevant for individuals carrying a single variant in AR genes. A growing literature has shown that carriers of AR disorders are exposed to an increased risk of complications in many diseases.^47,48^ Although few systematic investigations exists in IEI,^49^ incomplete penetrance and variable expressivity are frequent for many genes.^50^ Moreover, haploinsufficiency of several AR genes have been associated or suspected to be associated with clinical manifestations.^8^ Here, our purpose was not to identify patients with immune cytopenia having an underlying IEI. We rather used a genetic epidemiology approach to identify the rare variants associated with an increased risk of immune cytopenia and to quantify this risk. We showed that carrying a single LoF variant in an AR gene associated with IEI was a risk factor for both ITP and AIHA in the two cohorts analyzed. We did not identify an association with predicted deleterious missense variants. Although we report the largest genetic investigation in immune cytopenia, the rarity of the disease limited the power of analyses, and we cannot conclude on the lack of association for these variants. Moreover, among the several limitations of in silico tools is the limited capacity to discriminate putative LoF and gain-of-function missense variants. This difference is especially relevant in IEI given that several genes lead to IEI through a gain-of-function mechanism.^8^ Further studies are required to investigate the effect of these variants. LoF in *TET2* were the most frequent in both UKB and AoU and in both ITP and AIHA. Haploinsufficiency in *TET2* has been previously reported as associated with autoimmune lymphoproliferative syndrome-like syndrome.^51^ Our result expand the spectrum of *TET2* haploinsufficiency and show the penetrance of immune cytopenia is incomplete. Increased sample size of both control and cases cohorts is required to investigate the effect of LoF of other genes given their very low frequency.

Although the link between immune cytopenia and SLE is known for a long time, its genetic basis remained unexplored. SLE is a complex autoimmune disease with a substantial genetic susceptibility mediated by common variants. The SNP-based genetic heritability of SLE obtained by genome wide association studies is estimated to be ∼33%.^52^ SLE is genetically correlated with other autoimmune diseases.^53^ The SNP-based genetic heritability of immune cytopenia and the correlation with other autoimmune disease have never been studied due to the rarity of the disease. Our current sample size still prevents faithful estimation of h2. To overcome this limitation, we took advantage of the existing PGS_SLE_. We show that the common-variant mediated genetic predisposition to SLE also increases the risk of immune cytopenia.

One major finding of our study was that genetic contribution to immune cytopenia was substantial and larger than initially though in adults. Around one third to one half of individuals may carry a genetically mediated susceptibility. However, this estimate should be interpreted with caution. On one side, additional genetic susceptibility may have not been detected. Our analyses do not capture some variants such as missense IEI variants, structural variants, and low-frequency CH. As such, one could speculate that a genetic susceptibility may be present in a larger proportion of individuals with immune cytopenia. But on the other side, most individuals carrying a genetic variant will not develop immune cytopenia. The presence of genetic variant should be interpreted as causal as in monogenic disorders, but rather as a risk factor. Moreover, defining the threshold of high genetic susceptibility is challenging for PGS. As many other autoimmune diseases, the pathophysiology of immune cytopenia is complex, and its occurrence is not only mediated by genetic factors but also environmental factors. Future studies are required to analyse the contribution and interaction between genetic and environmental factors.

The second major finding of our analysis is the independence and quantification of the liability of the four types of genetic variants investigated. We showed that the genetic susceptibility is frequent but also heterogeneous. Somatic variants, especially hematological malignancies, were those associated with the largest risk increase. Our result suggest that an interaction may exist between hematological malignancies and clonal hematopoiesis. This would suggest these two types of somatic variants may contribute to immune cytopenia through at least partially similar mechanism. The risk carried by rare and common germline variants were similar and lower than somatic variants. Further studies with larger sample size may allow further discriminating the risk carried by subsets of germline and somatic variants.

Our results strongly support investigating the consequences of the different genetic variants on immune cytopenia phenotype. Both ITP and AIHA have highly heterogeneous course with some patients presenting refractory and/or chronic diseases.^54,55^ The underlying pathophysiology may differ across the spectrum of severity.^54,56,57^ Whether the different genetic variants lead to different endotypes with distinct pathophysiology warrants further investigation.

This comprehensive investigation of four different genetic variants can also pave the way of analyses in other autoimmune diseases. Common and rare variants are well known genetic risk factors of autoimmune diseases.^58–60^ Hematological malignancies and clonal hematopoiesis have been associated with various autoimmune diseases.^61–63^ Our analyses provide a framework for future investigations deciphering the respective role in other autoimmune diseases.

The joint analyses of the four types of variants allowed stratifying the risk of immune cytopenia in the general population. Although immune cytopenia remains a rare event, the different groups identified carried markedly different cumulative incidences. This stratification provides a unique tool to refine the risk of immune cytopenia in an individual. Of note, given that somatic variants may appear throughout life, the risk of an individual may evolve over time. Strikingly, we also showed that germline rare and common genetic variants modulate the risk of immune cytopenia in individuals carrying somatic variants. This increased risk may be clinically relevant in the assessment of hematological malignancies- associated complications to guide the choice of treatment. This is especially the case while considering using immune checkpoint inhibitors, which carry a substantial risk of immune cytopenia.^64^ Given the expansion of such approaches, assessing germline genetic variants may allow stratifying the risk of immune cytopenia.

Several limitations should be considered in the interpretation of our results. As many other genetic epidemiology studies, we face the limitations of biobanks in term of representativity and data accuracy. As ITP is a diagnosis of exclusion, misdiagnosis can concern a substantial part of the individuals.^65,66^ Although this is unlikely to lead to false positive results, especially given the consistency of our findings with AIHA, this could lead to underestimate the associations and to false negative results. Biobanks also provide limited follow-up data relevant to immune cytopenia such as the subtype of AIHA and the clinical course. UKB and AoU have distinct structure. The concordance of our results in the two cohorts strengthen our results. Nevertheless, the difference in term of population can have affected the accuracy of our estimates.

In sum, we provide the first estimation of the genetic susceptibility liability in immune cytopenia. A substantial proportion of individuals carried a susceptibility variant, with heterogeneous underlying variants. These variants allowed stratifying the risk of immune cytopenia. These results pave the way toward disentangling immune cytopenia heterogeneity and personalized risk estimation and management.

## Data Availability

The data that support the findings of this study are available from the UK Biobank and All of Us Program. Restrictions apply to the availability of these data, which were used under license for this study. Data are available from https://www.ukbiobank.ac.uk/ and https://www.researchallofus.org/ with permission from the UK Biobank and All of Us Program. This research has been conducted using the UK Biobank Resource under Application Number 399545.
Code used for this study is available in github (https://github.com/steniorj/genetic_contributors_aic).

https://github.com/steniorj/genetic_contributors_aic

## Ethic statement

Approval was granted by the CHU Sainte-Justine Research Ethic Board. The research was conducted according to the declaration of Helsinki.

## Data sharing statement

The data that support the findings of this study are available from the UK Biobank and All of Us Program. Restrictions apply to the availability of these data, which were used under license for this study. Data are available from https://www.ukbiobank.ac.uk/ and https://www.researchallofus.org/ with permission from the UK Biobank and All of Us Program. This research has been conducted using the UK Biobank Resource under Application Number 399545.

Code used for this study is available in github (https://github.com/steniorj/genetic_contributors_aic).

## Author Contribution

S.D.S.F. and E.L. performed the analyses. J.B. and R.M. created the figures. M.A.L. provided analytical insights. T.P. initiated and supervised the study. S.D.S.F. and T.P. wrote the first draft. All the authors approved the final version of the manuscript.

