## Supplemental data for "Rare and Common Germline and Somatic Variants Shape Immune Cytopenia Risk and Enable Risk Stratification"

| Supplemental Table | Page |
| --- | --- |
| Supplemental Table 1. Cohort description | 2 |
| Supplemental Table 2. Rare variant burden analysis | 4 |
| Supplemental Table 3. By-gene rare variant burden analysis. | 5 |
| Supplemental Table 4. Association between PGS_SLE_ and immune cytopenia. | 6 |
| Supplemental Table 5. Respective contribution of LoF variants, PGS_SLE_ and hematologic malignancies to immune cytopenia. | 7 |
| Supplemental Table 6. Risk stratification according to LoF variants, PGS_SLE_ and hematologic malignancies. | 7 |
| Supplemental Table 7. Association between LoF variants, PGS_SLE_, hematologic malignancies and clonal hematopoiesis with immune cytopenia. | 8 |
| Supplemental Table 8. Risk stratification according to LoF variants, PGS_SLE_, clonal hematopoiesis, and hematologic malignancies. | 9 |
| Supplemental Table 9. Interaction between LoF variants, PGS_SLE_, hematologic malignancies and clonal hematopoiesis with immune cytopenia. | 10 |
| Supplemental Table 10. Association between PGS_SLE_, LoF variants, hematologic malignancies and clonal hematopoiesis with age at immune cytopenia diagnosis. | 11 |
| Supplemental Table 11. Association between PGS_SLE_ and LoF variants with immune cytopenia in high-risk individuals. | 12 |
| Supplemental Table 12. LoF and PGS_SLE_ allow stratification of immune cytopenia risk among two-high-risk populations. | 12 |

**Supplemental Table 1. Cohort description**

| Diagnostic | ICD10 code | Description | Number of patients  UKB/AoU | Mean age (± SD) at diagnostic  UKB/AoU |
| --- | --- | --- | --- | --- |
| ITP | D69.3 | Idiopathic thrombocytopenic purpura | 937 / 802 | 64.35 (± 10.55) / 56.13 (± 17.03) |
| AIHA | D59.1 | Other autoimmune hemolytic anemias | 251 / 236 | 66.30 (± 9.43) / 57.72 (± 17.41) |
| ITP and AIHA | D69.3;  D59.1 | Idiopathic thrombocytopenic purpura;  Other autoimmune hemolytic anemias | 24 / 22 | 60.81 (± 10.63) / 49.38 (± 20.58) |
| SLE | M32 | Systemic lupus erythematosus (SLE) | 802 / 3122 | 59.72 (± 11.29) / 48.91 (± 14.56) |
| Hematological malignancies | C91.1  C81; | Chronic lymphocytic leukemia of B-cell type  Hodgkin lymphoma; | 8949 / 3534 | 64.19 (± 11.03) / 61.74 (± 14.68) |
|  | C82; | Follicular lymphoma; |  |  |
|  | C83; | Non-follicular lymphoma; |  |  |
|  | C84; | Mature T/NK-cell lymphomas; |  |  |
|  | C85; | Other and unspecified types of non-Hodgkin lymphoma; |  |  |
|  | C86; | Other specified types of T/NK-cell lymphoma; |  |  |
|  | C88; | Malignant immunoproliferative diseases; |  |  |
|  | C88.4 | Extranodal marginal zone B-cell lymphoma of mucosa-associated lymphoid tissue [MALT-lymphoma] |  |  |
|  | C90.1; | Plasma cell leukemia; |  |  |
|  | C91.0; | Acute lymphoblastic leukemia; |  |  |
|  | C91.3; | Prolymphocytic leukemia of B-cell type; |  |  |
|  | C91.4; | Hairy-cell leukemia; |  |  |
|  | C91.5; | Adult T-cell lymphoma/leukemia [HTLV-1-associated]; |  |  |
|  | C91.6; | Prolymphocytic leukemia of T-cell type; |  |  |
|  | C91.7; | Other lymphoid leukemia; |  |  |
|  | C91.8; | Mature B-cell leukemia Burkitt-type; |  |  |
|  | C91.9; | Lymphoid leukemia, unspecified; |  |  |
|  | C92; | Myeloid leukemia; |  |  |
|  | C93; | Monocytic leukemia; |  |  |
|  | C94; | Other leukemias of specified cell type; |  |  |
|  | C95; | Leukemia of unspecified cell type; |  |  |
|  | D47.1; | Chronic myeloproliferative disease; |  |  |
|  | D47.5; | Chronic eosinophilic leukemia [hypereosinophilic syndrome]; |  |  |
|  | C96.7; | Other specified malignant neoplasms of lymphoid, hematopoietic and related tissue; |  |  |
|  | C96.9; | Malignant neoplasms of lymphoid, hematopoietic and related tissue, unspecified; |  |  |
|  | D47.4; | Osteomyelofibrosis; |  |  |
|  | D47.7; | Other specified neoplasms of uncertain or unknown behavior of lymphoid, hematopoietic and related tissue; |  |  |
|  | D47.9; | Neoplasm of uncertain or unknown behavior of lymphoid, hematopoietic and related tissue, unspecified; |  |  |
|  | D75.2; | Essential thrombocytosis; |  |  |
|  | M47.17; | Other spondylosis with myelopathy (Lumbosacral region); |  |  |
|  | Z85.7; | Personal history of other malignant neoplasms of lymphoid, hematopoietic and related tissues |  |  |
| Clonal Hematopoiesis | - | - | 15,613 | 61.28 (± 6.66) |

**Supplemental Table 2. Rare variant burden analysis.** We considered rare damaging variants in 62 genes associated with ITP and/or AIHA. We performed multivariate logistic regression adjusted on age at last follow-up, sex, center for AoU cohort, and the first 10 principal components. The gene set of AR genes included 39 genes

|  | ITP and/or AIHA | | | |  | ITP | | | |  | AIHA | | | |
| --- | --- | --- | --- | --- | --- | --- | --- | --- | --- | --- | --- | --- | --- | --- |
|  | aOR | 95% CI | | p |  | aOR | 95% CI | | p |  | aOR | 95% CI | | p |
|  |  | Low | High |  |  |  | Low | High |  |  |  | Low | High |  |
| UK Biobank, MAF <0.1% | | | | | | | | | | | | | | |
| LoF | 2.26 | 1.58 | 3.11 | 1.86x10^-6^ |  | 2.23 | 1.49 | 3.17 | 2.96x10^-5^ |  | 2.76 | 1.33 | 4.93 | 0.002 |
| REVEL 0.5 | 1.03 | 0.86 | 1.23 | 0.74 |  | 0.99 | 0.80 | 1.22 | 0.98 |  | 1.20 | 0.80 | 1.70 | 0.34 |
| REVEL 0.9 | 1.24 | 0.62 | 2.17 | 0.51 |  | 1.22 | 0.55 | 2.28 | 0.57 |  | 1.76 | 0.43 | 4.57 | 0.33 |
| LoF + REVEL 0.9 | 1.91 | 1.40 | 2.53 | 1.91x10^-5^ |  | 1.88 | 1.32 | 2.58 | 1.92x10^-4^ |  | 2.42 | 1.30 | 4.06 | 0.002 |
| AlphaMissense | 1.02 | 0.84 | 1.23 | 0.78 |  | 0.96 | 0.77 | 1.19 | 0.76 |  | 1.29 | 0.87 | 1.81 | 0.174 |
| LoF, AR only genes | 2.05 | 1.38 | 2.91 | 1.36x10^-4^ |  | 2.31 | 1.52 | 3.33 | 2.79x10^-5^ |  | 1.28 | 0.39 | 2.96 | 0.62 |
| UK Biobank, MAF <1% | | | | | | | | | | | | | | |
| LoF | 1.82 | 1.28 | 2.49 | 4.43x10^-4^ |  | 1.80 | 1.21 | 2.55 | 0.001 |  | 2.18 | 1.05 | 3.93 | 0.01 |
| REVEL 0.5 | 1.14 | 1.00 | 1.30 | 0.04 |  | 1.150 | 0.990 | 1.320 | 0.06 |  | 1.160 | 0.86 | 1.51 | 0.30 |
| REVEL 0.9 | 1.15 | 0.75 | 1.65 | 0.48 |  | 1.210 | 0.769 | 1.790 | 0.37 |  | 1.210 | 0.47 | 2.47 | 0.64 |
| LoF + REVEL 0.9 | 1.46 | 1.12 | 1.86 | 0.003 |  | 1.49 | 1.11 | 1.94 | 0.005 |  | 1.660 | 0.95 | 2.66 | 0.05 |
| All of Us, MAF <0.1% | | | | | | | | | | | | | | |
| LoF | 1.44 | 1.08 | 1.91 | 0.01 |  | 1.54 | 1.01 | 2.38 | 0.04 |  | 2.22 | 1.14 | 4.33 | 0.02 |
| LoF, AR only genes | 1.62 | 1.19 | 2.20 | 0.002 |  | 1.78 | 1.13 | 2.81 | 0.01 |  | 2.54 | 1.25 | 5.15 | 0.01 |

**Supplemental Table 3. By-gene rare variant burden analysis.** We considered rare LoF variants found in any of the 62 genes associated with ITP and/or AIHA. For each immune cytopenia, we considered only genes with at least one variant in cases. We performed multivariate logistic regression adjusted on age at last follow-up, sex, center for AoU cohort, and the first 10 principal components. Each gene was tested in a different multivariate model.

|  |  | ITP | | | |  | AIHA | | | |
| --- | --- | --- | --- | --- | --- | --- | --- | --- | --- | --- |
|  |  | aOR | 95% CI | | FDR |  | aOR | 95% CI | | FDR |
|  |  |  | Low | High |  |  |  | Low | High |  |
| UK Biobank |  |  |  |  |  |  |  |  |  |  |
| *TNFRSF13B* |  | 1.44 | 0.20 | 10.30 | 0.994 |  | 5.23 | 0.73 | 37.56 | 0.723 |
| *NCF1* |  |  |  |  |  |  | 8.18 | 1.07 | 62.20 | 0.327 |
| *PIK3CG* |  | 2.44 | 0.34 | 17.43 | 0.994 |  |  |  |  |  |
| *ATM* |  | 1.73 | 0.56 | 5.34 | 0.994 |  |  |  |  |  |
| *IKBKB* |  | 3.97 | 0.55 | 28.48 | 0.994 |  |  |  |  |  |
| *PNP* |  | 4.81 | 0.67 | 34.50 | 0.798 |  |  |  |  |  |
| *ICOS* |  | 7.90 | 1.09 | 57.08 | 0.327 |  |  |  |  |  |
| *KMT2D* |  | 9.15 | 1.27 | 66.11 | 0.255 |  | 32.07 | 4.42 | 232.65 | 0.011 |
| *RAG1* |  | 4.16 | 1.33 | 12.97 | 0.139 |  | 10.15 | 2.51 | 40.97 | 0.018 |
| *CYBA* |  | 12.50 | 1.72 | 90.96 | 0.136 |  |  |  |  |  |
| *DCLRE1C* |  | 6.23 | 1.54 | 25.18 | 0.123 |  |  |  |  |  |
| *MALT1* |  | 13.72 | 3.36 | 56.08 | 5.74x10^-3^ |  |  |  |  |  |
| *LCP2* |  |  |  |  |  |  | 19.71 | 4.86 | 79.90 | 8.02x10^-4^ |
| *FASLG* |  | 178.48 | 18.51 | 1720.70 | 2.63x10^-4^ |  | 635.73 | 64.94 | 6223.51 | 1.58x10^-6^ |
| *TET2* |  | 9.66 | 5.41 | 17.26 | 2.03x10^-12^ |  | 7.41 | 2.10 | 26.18 | 0.025 |
| All of Us |  |  |  |  |  |  |  |  |  |  |
| *TET2* |  | 3.00 | 1.24 | 7.28 | 0.01 |  | 9.70 | 3.95 | 16.01 | 0.05 |

**Supplemental Table 4. Association between PGS_SLE_ and immune cytopenia.** We performed multivariate logistic regression with three different standardized PGS. We adjusted for LoF and excluded individuals with SLE in some analyses.

|  | ITP and/or AIHA | | | |  | ITP | | | |  | AIHA | | | |
| --- | --- | --- | --- | --- | --- | --- | --- | --- | --- | --- | --- | --- | --- | --- |
|  | aOR | 95% CI | | p |  | aOR | 95% CI | | p |  | aOR | 95% CI | | p |
|  |  | Low | High |  |  |  | Low | High |  |  |  | Low | High |  |
| UK Biobank | | | | | | | | | | | | | | |
| Standard PGS | 1.19 | 1.12 | 1.26 | 1.6x10^-8^ |  | 1.13 | 1.05 | 1.20 | 5.2x10^-4^ |  | 1.19 | 1.12 | 1.26 | 1.5x10^-8^ |
| Standard PGS adjusted for LoF | 1.16 | 1.09 | 1.23 | 6.5x10^-7^ |  | 1.10 | 1.03 | 1.18 | 0.003 |  | 1.43 | 1.27 | 1.62 | 1.2x10^-8^ |
| Standard PGS  after exclusion of patients with ITP/AIHA and SLE | 1.17 | 1.10 | 1.24 | 6.9x10^-7^ |  | 1.11 | 1.04 | 1.19 | 0.002 |  | 1.42 | 1.25 | 1.61 | 1.3x10^-7^ |
| All of Us | | | | | | | | | | | | | | |
| PGS000196 | 1.17 | 1.11 | 1.23 | 5.1x10^-9^ |  | 1.16 | 1.09 | 1.23 | 9.4x10^-7^ |  | 1.24 | 1.11 | 1.38 | 7.5x10^-5^ |
| PGS004917 | 1.11 | 1.07 | 1.15 | 7.4x10^-10^ |  | 1.10 | 1.06 | 1.14 | 1.5x10^-6^ |  | 1.19 | 1.11 | 1.28 | 4.5x10^-7^ |
| PGS000196  adjusted for LoF | 1.10 | 1.05 | 1.16 | 6.9x10^-5^ |  | 1.08 | 1.03 | 1.14 | 0.003 |  | 1.22 | 1.10 | 1.34 | 8.8x10^-5^ |
| PGS004917  adjusted for LoF | 1.07 | 1.03 | 1.10 | 3.9x10^-5^ |  | 1.05 | 1.01 | 1.09 | 0.006 |  | 1.17 | 1.10 | 1.25 | 1.1x10^-6^ |
| PGS000196  after exclusion of patients with ITP/AIHA and SLE | 1.11 | 1.05 | 1.18 | 2.5X10^-4^ |  | 1.09 | 1.02 | 1.16 | 0.008 |  | 1.21 | 1.08 | 1.35 | 0.001 |
| PGS004917  after exclusion of patients with ITP/AIHA and SLE | 1.08 | 1.04 | 1.12 | 1.9x10^-5^ |  | 1.06 | 1.02 | 1.11 | 0.003 |  | 1.17 | 1.09 | 1.26 | 3.0x10^-5^ |

**Supplemental Table 5. Respective contribution of LoF variants, PGS_SLE_ and hematologic malignancies to immune cytopenia.** We performed multivariate logistic and Cox regression with LoF variants in IEI genes, standardized PGS_SLE_ (Standard PGS in the UKB and PGS004917 in AoU), and covariates. Hematologic malignancies having occurred after immune cytopenia were not considered. For Cox regression, we used inclusion in the UKB and first clinical record in AoU as initial timepoints of analyses.

|  | UK Biobank | | | |  | All of Us | | | |
| --- | --- | --- | --- | --- | --- | --- | --- | --- | --- |
|  | aOR/aHR | 95% CI | | p |  | aOR/aHR | 95% CI | | p |
|  |  | Low | High |  |  |  | Low | High |  |
| Logistic regression | | | | | | | | | |
| LoF variants | 1.99 | 1.39 | 2.74 | 6.73x10^-5^ |  | 1.41 | 1.02 | 1.78 | 0.012 |
| PGS_SLE_ | 1.17 | 1.10 | 1.24 | 2.08x10^-07^ |  | 1.11 | 1.07 | 1.15 | 8.42x10^-10^ |
| Hematologic malignancies | 14.26 | 10.8 | 14.6 | 1.36x10^-238^ |  | 4.68 | 3.60 | 5.99 | 1.24x10^-32^ |
| Cox regression | | | | | | | | | |
| LoF variants | 1.84 | 1.25 | 2.72 | 0.002 |  | 1.38 | 1.02 | 1.85 | 0.032 |
| PGS_SLE_ | 1.15 | 1.07 | 1.23 | 2.90x10^-05^ |  | 1.11 | 1.07 | 1.14 | 1.51x10^-9^ |
| Hematologic malignancies | 14.24 | 12.11 | 16.74 | 3.80x10^-227^ |  | 3.86 | 2.99 | 4.99 | 2.26x10^-25^ |

**Supplemental Table 6. Risk stratification according to LoF variants, PGS_SLE_ and hematologic malignancies.** We stratified the population in three groups based on the genetic variants carried and performed Cox regression multivariate adjusted for age, sex and 10 principal components. We used inclusion in the UKB and first clinical record in AoU as initial timepoints of analyses.

|  | UK Biobank | | | |  | All of Us | | | |
| --- | --- | --- | --- | --- | --- | --- | --- | --- | --- |
| Risk group | aHR | 95% CI | | p |  | aHR | 95% CI | | p |
|  |  | Low | High |  |  |  | Low | High |  |
| Considering PGS_SLE_ >1 SD | | | | | | | | | |
| Hematologic malignancies compared to no variants | 16.67 | 14.29 | 20.00 | 2.05x10^-228^ |  | 5.00 | 3.85 | 6.25 | 7.59x10^-36^ |
| LoF or PGS_SLE_ >1 SD compared to no variants | 1.37 | 1.14 | 1.660 | 6.46x10^-4^ |  | 1.62 | 1.38 | 1.90 | 4.05x10^-9^ |
| Hematologic malignancies compared to LoF or PGS_SLE_ >1 SD | 12.50 | 9.09 | 14.29 | 4.29x10^-105^ |  | 3.03 | 2.54 | 4.42 | 1.13x10^-14^ |
| Considering PGS_SLE_ >1.5 SD | | | | | | | | | |
| Hematologic malignancies compared to no variants | 16.67 | 14.28 | 20 | 5.35x10^-233^ |  | 4.76 | 3.70 | 6.25 | 3.17x10^-34^ |
| LoF or PGS_SLE_ >1.5 SD compared to no variants | 1.64 | 1.30 | 2.05 | 1.85x10^-05^ |  | 1.50 | 1.22 | 1.84 | 1.21x10^-4^ |
| Hematologic malignancies compared to LoF or PGS_SLE_ >1.5 SD | 10 | 7.69 | 12.5 | 1.17x10^-65^ |  | 3.12 | 2.72 | 4.34 | 9.06x10^-3^ |
| Considering PGS_SLE_ >2 SD | | | | | | | | | |
| Hematologic malignancies compared to no variants | 16.67 | 14.28 | 20 | 9.24x10^-233^ |  | 4.76 | 3.57 | 5.88 | 8.67x10^-34^ |
| LoF or PGS_SLE_ >2 SD compared to no variants | 1.70 | 1.25 | 2.30 | 6.68x10^-4^ |  | 1.55 | 1.19 | 2.02 | 1.03x10^-3^ |
| Hematologic malignancies compared to LoF or PGS_SLE_ >2 SD | 0.09 | 6.67 | 12.5 | 2.35x10^-38^ |  | 2.94 | 2.08 | 4.16 | 1.53x10^-9^ |

**Supplemental Table 7. Association between LoF variants, PGS_SLE_, hematologic malignancies and clonal hematopoiesis with immune cytopenia.** We performed multivariate Cox regression with LoF variants, standardized Standard PGS_SLE_ and covariates. Hematologic malignancies and clonal hematopoiesis having occurred after immune cytopenia were not considered. We used inclusion in the UKB as initial timepoint of analyses.

|  | UK Biobank | | | |
| --- | --- | --- | --- | --- |
|  | aHR | 95% CI | | p |
|  |  | Low | High |  |
| Hematologic malignancies | 13.60 | 11.56 | 16.01 | 5.9x10^-216^ |
| Clonal hematopoiesis | 2.14 | 1.70 | 2.69 | 6.6x10^-11^ |
| LoF variants | 1.67 | 1.36 | 2.47 | 0.009 |
| PGS_SLE_ | 1.15 | 1.08 | 1.24 | 2.2x10^-5^ |

**Supplemental Table 8. Risk stratification according to LoF variants, PGS_SLE_, clonal hematopoiesis, and hematologic malignancies.** We stratified the population in four groups based on the genetic variants carried and performed Cox regression multivariate adjusted for age, sex and 10 principal components. We used inclusion in the UKB as initial timepoint of analyses.

|  | UK Biobank | | | |
| --- | --- | --- | --- | --- |
| Risk group | aHR | 95% CI | | p |
|  |  | Low | High |  |
| Considering PGS_SLE_ >1 SD | | | | |
| Hematologic malignancies compared to no variants | 16.67 | 14.29 | 20.00 | 2.05x10^-233^ |
| Clonal hematopoiesis compared to no variants | 2.85 | 2.08 | 3.84 | 7.23x10^-12^ |
| LoF or PGS_SLE_ >1 SD compared to no variants | 1.47 | 1.21 | 1.78 | 5.46x10^-5^ |
| Hematologic malignancies compared to LoF or PGS_SLE_ >1 SD | 12.50 | 9.09 | 14.29 | 1.27x10^-105^ |
| Hematologic malignancies compared to clonal hematopoiesis | 6.25 | 4.34 | 8.33 | 3.73x10^-29^ |
| Clonal hematopoiesis compared to LoF or PGS_SLE_ >1 SD | 1.89 | 1.37 | 2.62 | 1.05x10^-4^ |
| Considering PGS_SLE_ >1.5 SD | | | | |
| Hematologic malignancies compared to no variants | 16.67 | 14.28 | 20 | 1.41x10^-237^ |
| Clonal hematopoiesis compared to no variants | 2.70 | 2 | 3.57 | 2.07x10^-5^ |
| LoF or PGS_SLE_ >1.5 SD compared to no variants | 1.75 | 1.38 | 2.22 | 1.98x10^-6^ |
| Hematologic malignancies compared to LoF or PGS_SLE_ >1.5 SD | 10 | 7.69 | 12.5 | 5.00x10^-66^ |
| Hematologic malignancies compared to clonal hematopoiesis | 6.25 | 4.54 | 8.33 | 9.58x10^-32^ |
| Clonal hematopoiesis compared to LoF or PGS_SLE_ >1.5 SD | 1.51 | 1.07 | 2.14 | 1.71x10^-2^ |
| Considering PGS_SLE_ >2 SD |  |  |  |  |
| Hematologic malignancies compared to no variants | 16.67 | 14.29 | 20.00 | 3.90x10^-237^ |
| Clonal hematopoiesis compared to no variants | 2.50 | 1.88 | 3.33 | 1.83X10^-10^ |
| LoF or PGS_SLE_ >2 SD compared to no variants | 1.81 | 1.22 | 2.43 | 1.90x10^-4^ |
| Hematologic malignancies compared to LoF or PGS_SLE_ >2 SD | 9.09 | 6.67 | 12.50 | 1.51x10^-38^ |
| Hematologic malignancies compared to clonal hematopoiesis | 6.67 | 4.76 | 9.09 | 3.61x10^-33^ |
| Clonal hematopoiesis compared to LoF or PGS_SLE_ >2 SD | 1.39 | 0.93 | 2.07 | 0.105 |

**Supplemental Table 9. Interaction between LoF variants, PGS_SLE_, hematologic malignancies and clonal hematopoiesis with immune cytopenia.** We performed separate multivariate analyses to test the interaction between genetic variants two by two, given our limited power to test all interactions simultaneously. All models included the four genetic variants, covariates, and one interaction.

|  | LoF variants | PGS_SLE_ | Hematologic malignancies |
| --- | --- | --- | --- |
|  | p-value | | |
| PGS_SLE_ | 0.11 | - | - |
| Hematologic malignancies | 0.71 | 0.40 | - |
| Clonal hematopoiesis | 0.09 | 0.08 | 0.01 |

**Supplemental Table 10. Association between PGS_SLE_, LoF variants, hematologic malignancies and clonal hematopoiesis with age at immune cytopenia diagnosis.** We performed in the UKB a single multivariate analysis with LoF variants, standardized PGS_SLE,_ hematologic malignancies, clonal hematopoiesis, and covariates in the UKB.

|  |  | UK Biobank | | | |
| --- | --- | --- | --- | --- | --- |
|  |  | aOR | 95% CI | | p |
|  |  |  | Low | High |  |
| LoF variants |  | 0.21 | 0.23 | 1.97 | 0.174 |
| PGS_SLE_ |  | 0.82 | 0.56 | 1.19 | 0.303 |
| Hematologic malignancies |  | 6.90 | 2.65 | 18.0 | 8.12x10^-5^ |
| Clonal hematopoiesis |  | 2.21 | 0.55 | 8.76 | 0.261 |

**Supplemental Table 11. Association between PGS_SLE_ and LoF variants with immune cytopenia in high-risk individuals.** We performed in the UKB multivariate analysis with LoF variants, standardized standard PGS_SLE_ and covariates. We analyzed individuals with hematologic malignancy and individuals with clonal hematopoiesis.

|  |  | LoF | | | |  | PGS_SLE_ | | | |
| --- | --- | --- | --- | --- | --- | --- | --- | --- | --- | --- |
|  |  | aOR | 95% CI | | p |  | aOR | 95% CI | | p |
|  |  |  | Low | High |  |  |  | Low | High |  |
| UK Biobank | | | | | | | | | | |
| Hematologic malignancies |  | 2.69 | 1.65 | 4.10 | 1.79x10^-5^ |  | 1.15 | 1.02 | 1.30 | 0.017 |
| Clonal hematopoiesis |  | 4.43 | 2.29 | 7.70 | 1.08x10^-6^ |  | 0.97 | 0.77 | 1.21 | 0.801 |

**Supplemental Table 12. LoF and PGS_SLE_ allow stratification of immune cytopenia risk among two-high-risk populations.** We performed multivariate analysis in individuals with hematologic malignancies according to the presence of LoF variants and/or PGS_SLE_ value > 1.5 SD and in individuals with clonal hematopoiesis according to the presence of LoF variantss

|  |  | Hematologic malignancies | | | |  | Clonal hematopoiesis | | | |
| --- | --- | --- | --- | --- | --- | --- | --- | --- | --- | --- |
|  |  | aHR | 95% CI | | p |  | aHR | 95% CI | | p |
|  |  |  | Low | High |  |  |  | Low | High |  |
| LoF variants and/or PGS_SLE_ value > 1.5 SD |  | 1.83 | 1.25 | 2.68 | 0.001 |  |  |  |  |  |
| LoF only |  |  |  |  |  |  | 4.37 | 2.43 | 7.84 | 6.3x10^-6^ |
